# Sources and pathways by which low-grade inflammation contributes to anaemia in rural African children from 6 months to 3 years of age: study protocol for observational studies IDeA 1 and IDeA 2

**DOI:** 10.1101/2024.02.09.24301930

**Authors:** Elizabeth Ledger, Hans Verhoef, Amadou T Jallow, Nicole Cunningham, Andrew M. Prentice, Carla Cerami

## Abstract

**Background:** Recent work suggests that persistent inflammation, even at low levels, could be more important than low dietary iron intake in the aetiology of iron deficiency and iron deficiency anaemia (IDA) in young children living in poor environments.

**Methods:** We will conduct 2 parallel observational studies in well and unwell rural Gambian children to identify the origins of chronic low-grade inflammation and characterise its relationship to iron handling and iron deficiency anaemia. IDeA Study 1 will enrol 120 well children attending our regular paediatric well-child clinics at 6, 12 and 18 months of age. IDeA Study 2 will enrol 200 sick children suffering from upper-respiratory tract infection, lower respiratory tract infection, gastroenteritis or urinary tract infection and study them on Days 0, 3, 7 and 14 after initial presentation. At each visit, children will be assessed for signs of inflammation. Full blood count and iron-related biomarkers (serum ferritin, serum iron, unsaturated iron-binding capacity, soluble transferrin receptor, transferrin) will be measured before and after an oral dose of ferrous iron to assess status and acute iron absorption. Inflammatory markers (C-reactive protein and *α*_1_-acid glycoprotein), hepcidin, erythroferrone and erythropoietin will be measured to characterize the anaemia of inflammation in these children.

**Conclusion:** We will assess the impact of acute and chronic low-grade inflammation on iron absorption and investigate the hypothesis that chronic inflammation, juxtaposed on a poor diet, causes a complex anaemia of inflammation which exacerbates iron deficiency by blocking both non-haem iron absorption and iron utilization by the bone marrow.

## BACKGROUND

Iron deficiency (ID) is the most prevalent nutritional deficiency worldwide and, with its consequent iron deficiency anaemia (IDA), is most prevalent in Sub-Saharan Africa and South Asia [1]. The Global Burden of Disease estimates that 1.225 billion people suffer from IDA and the consequent years lived with disability (YLDs) outweigh all other nutritional deficiencies, haemoglobinopathies and haemolytic anaemias combined [1–2]. A 50% reduction in anaemia rates in women of reproductive age is one of WHO’s 6 key Global Targets for 2025. However, it has been estimated that at current rates of progress this target will not be reached for 100 years. Similar challenges exist for the rates of anaemia in children.

### Effects of IDA

Iron deficiency is a strong risk factor for adverse pregnancy outcomes including maternal mortality. During foetal and child development, iron is required for diverse aspects of neural formation, and deficiency is associated with delayed maturation of the brain and impaired cognition [3,4]. Importantly, deficits occurring in these early critical windows leave children with permanent damage that affects educational potential and hence human capital [5]. Iron deficiency may also impair immune responses, especially cellular immunity, and hence may limit the effectiveness of early childhood vaccines [6]. Interventions to correct ID face two interlocking challenges; potential hazard and lack of efficacy.

### Potential hazards of iron supplementation

The possible hazards of iron administration in LMICs arise from the fact that iron stimulates the growth of many bacteria, protozoa and fungi [7]. Large epidemiological studies have shown that ID protects against malaria and several RCTs have shown that administration of iron can increase the risk of serious adverse outcomes in children (malaria, severe diarrhoea and respiratory tract infections) leading to excess hospitalisations and deaths [8]. We have recently described the most likely mechanism of how IDA protects against malaria, and how iron supplementation enhances the risk [9,10]. We have demonstrated how acute iron administration enhances bacterial growth rates [11,12]. These risks are driven at least in part by the use of non-physiological high doses of readily absorbable iron thought necessary to overcome the lack of efficacy.

### Efficacy of oral iron supplementation

The limited efficacy of iron administration is a universal problem across all iron supplementation trials in LMICs and especially Africa. For example, we have recently completed 2 trials in rural Gambia with 400 children aged 6-24m [13,14] and 500 second trimester pregnant women [12]. Despite administering iron for 12 weeks according to the WHO recommendations (12mg/d to children and 60mg/d to pregnant women) with an additional 15 micronutrients and with all doses taken under direct supervision, 45% of women at delivery and 75% of the children remained anaemic [12,14]. Recent meta-analysis of 14 studies with 3011 patients found that iron supplementation reduced the risk of anemia (defined as Hb <110g/dl) by 45% and increased mean haemoglobin by 0.6g/l [15].

### The role of hepcidin

The discovery of the hormone hepcidin, a largely liver-derived peptide that acts as a master regulator of iron absorption and distribution, has yielded many new insights into the real-life biology of iron, iron-related infections and anemia [16]. Hepcidin is down regulated by iron deficiency and erythroid drive and up-regulated by iron sufficiency or overload and by inflammation [7,16]. Hepcidin blocks iron absorption and recycling by down regulating ferroportin on the surface of enterocytes and macrophages. Conversely low levels of circulating hepcidin promote duodenal absorption of dietary iron, and permit rapid recycling of iron from aged or damaged erythrocytes.

### Anaemia of inflammation

In patients with autoimmune disease, renal disease and cancer there is a well described pathway to anaemia of inflammation where pro-inflammatory cytokines and upregulation of hepcidin lead to a reduction in red cell production [17]. Our recent work has shown that persistent inflammation (even at surprisingly low levels) could be more important than low dietary iron in the aetiology of ID and IDA [18]. In the HIGH study [13] hepcidin and haemoglobin were measured weekly and haematological, inflammatory and iron biomarkers were measured at baseline, 7 weeks and 12 weeks in 407 anaemic (Hb <11g/dl) but otherwise healthy Gambian children (6-27 months). Each child maintained a remarkably constant hepcidin level and half consistently maintained levels that indicated physiological blockade of iron absorption. Hepcidin was strongly predicted by nurse-ascribed adverse events with dominant signals from respiratory infections and fevers [18]. In multiple linear regression analysis, serum C-reactive protein (CRP) was the dominant predictor of hepcidin and contributed to iron blockade even at very low levels. It was concluded that even low-grade inflammation was an important driver of IDA in rural African children.

Previous work has shown that iron incorporation was lower on Day 0 in children presenting with acute febrile malaria and on Day 15 of convalescence than in Hb-matched children with iron deficiency alone [19]. Subsequent analysis of the same data set revealed that hepcidin concentration best predicted iron incorporation [20]. In a study in Tanzanian children, hepcidin concentrations normalized by 4 weeks after treatment for malaria [21] and a recent study in Uganda found that children recovering from malaria incorporated more iron if they were given iron supplements starting 28 days after treatment for malaria than if given immediately [22]. The impact of other infections on iron absorption is unknown and requires clarity to guide clinical strategies for enhancing iron absorption.

We hypothesise that chronic inflammation (even at low levels), juxtaposed upon a poor diet, causes anaemia of inflammation [17] which, by diverse possible pathways, exacerbates iron deficiency by blocking both non-haem iron absorption and iron utilization by the bone marrow (see **Figure 1**).

**Figure 1:**
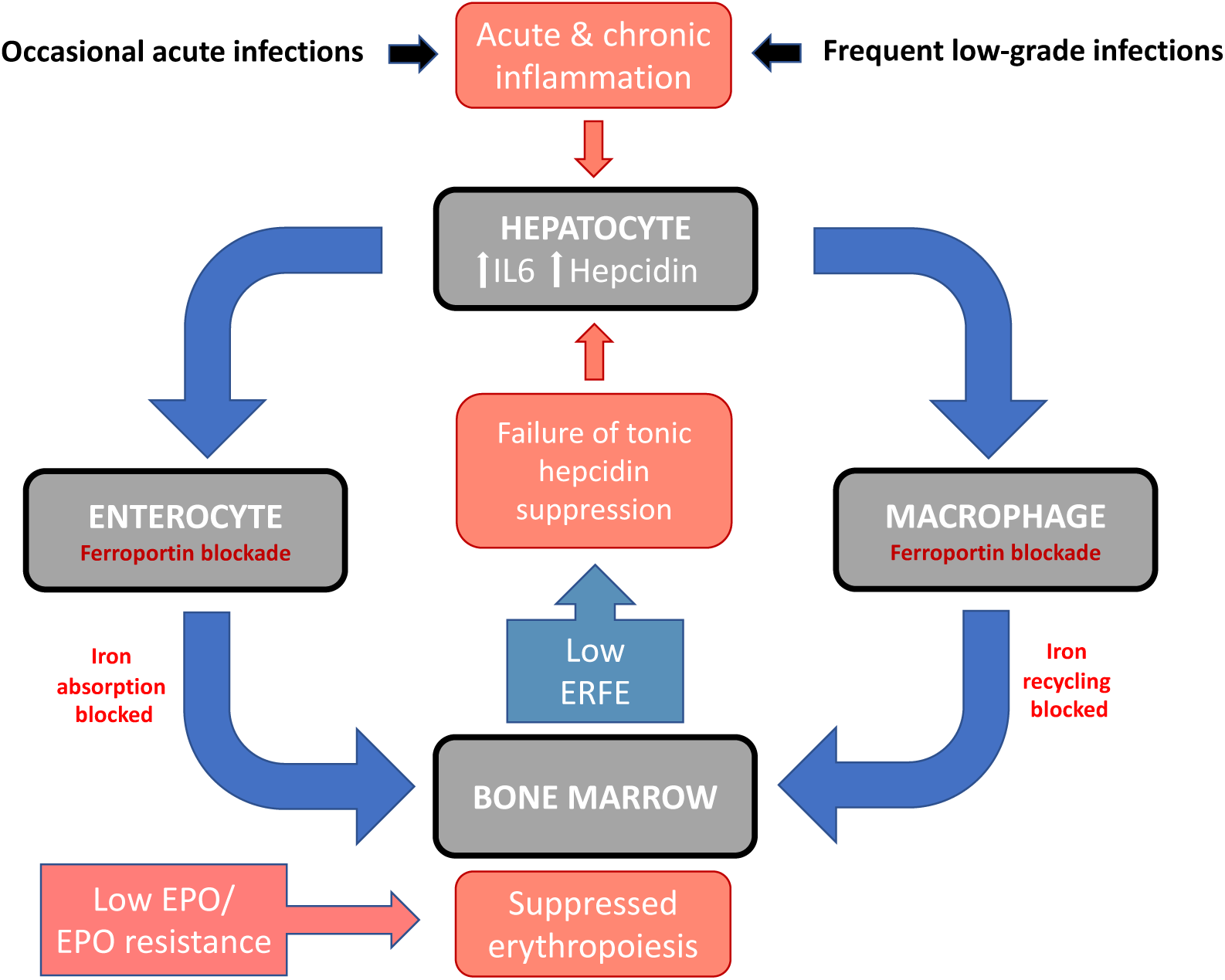
Pathways by which inflammation in young children contributes to an anaemia of inflammation. IL-6, interleukin-6 (and related inflammatory signals); ERFE, erythroferrone; EPO, erythropoietin

In our prior study it was not possible to define the source(s) of the low-grade chronic inflammation because the children did not have detailed clinical examinations. The purpose of the current studies is to locate the origin(s) of inflammation in apparently well and sick children. In the cohort of well children their inflammation presumably arises from sub-clinical silent insults such as skin infections, poor oral hygiene, gum disease, dental caries or gastrointestinal enteropathy. The sick children will be recruited into 4 diagnostic categories representing the most common causes of fever in this setting (now that the incidence of malaria has declined substantially); upper respiratory tract infections (URTI), lower respiratory tract infections (LRTI), gastroenteritis and urinary tract infections

### Study objectives

The overarching goal is to clarify the relationship between chronic low-grade inflammation and iron deficiency anaemia in rural African children. To this end the specific objectives are:

#### IDeA Study 1

<u>1)</u> To confirm our prior observations that rural Gambian children frequently display persistent inflammatory-mediated raised levels of hepcidin and to assess changes from 6-18 months of age;
<u>2)</u> To seek to establish the likely source(s) of the persistent low-grade inflammation in apparently well children living in rural Gambia between the ages of 6-18 months of age.

#### IdeA Study 2

1) To seek to establish the likely source(s) of inflammation in sick children presenting to our pediatric clinic in rural Gambia between 6-36 months of age with any of 4 diagnostic categories; upper respiratory tract infections (URTI), lower respiratory tract infections (LRTI), gastroenteritis or urinary tract infections (UTI).

#### Secondary objectives for both studies

1) To retest the existing hepcidin threshold of 5ng/ml (based on the Bachem ELISA; [23]) for discriminating iron absorbers from non-absorbers by repeating our prior ROC analysis based on a much larger sample;
2) To establish the role of inflammation in the relationship between erythropoietin concentration (EPO) and haematological concentration s;
3) To describe the interrelationships between serum markers of systemic inflammation (CRP/*α*_1_-acid glycoprotein, AGP) and their associations with serum concentrations of hepcidin and erythroferrone (ERFE);
4) To examine the influence of EPO on the association between haematological status and ERFE.

### Rationale supporting the secondary objectives

Secondary objective 1: Using ^57^Fe/^58^Fe tracer studies of dietary iron utilisation in rural Gambian children [18,19] we previously determined that a threshold level of hepcidin (at 5.5ng/ml using the BACHEM ELISA) distinguished good iron absorbers from poor iron absorbers [23]. Although the ROC AUC was high at 0.90 this analysis has two limitations: a) it was based on a very small sample; and b) half of the children were recovering from an acute episode of malaria. This analysis will now be repeated to validate (or adjust) the threshold based on a much larger sample size and data from well children. Importantly our new dataset will allow us to test whether the threshold varies by age, anaemia status and at different levels of inflammation. Note that the threshold can be adjusted for use with other hepcidin assays [24].

Secondary objective 2: We hypothesize that anaemic children with low-grade inflammation have an anaemia of inflammation with concomitant iron deficiency that results in inappropriately low levels of EPO production and reticulocytosis for their erythroid mass. Lack of responsiveness to EPO is an important component and has been previously described in acute and convalescent malaria in African children with high levels of inflammation [eg 25]. We will explore the relationship between persistent lower grade inflammation and the production of EPO in desponse to anaemia. We will achieve this by establishing the role of inflammation in the relationship between EPO and haemoglobin (as a measure of erythroid mass relative to total blood volume).

Secondary objectives 3 and 4: Erythroferrone is the recently-discovered hormone that mediates hepcidin suppression during stress erythropoiesis [26,27]. First, we will conduct a hypothesis-free exploratory analysis to assess whether ERFE behaves as predicted based upon mouse models (i.e. is up-regulated by stress erythropoiesis and inversely related to hepcidin). As summarised in **Fig 1** we hypothesise that there may be a vicious cycle initiated by inflammation and then perpetuated by the consequent low levels of (iron-restricted) erythropoiesis, leading to low ERFE and loss of hepcidin suppression.

## SUBJECTS AND METHODS

### Study setting

MRC The Gambia Keneba field station is situated in rural West Africa where there are dramatic seasonal shifts from wet to dry seasons and where the majority of the population are subsistence farmers. The field station runs a well-accessed free primary health care facility and also is the home of the Keneba Biobank Project which contains blood samples from consenting individuals in 36 villages with 96% participation [28]. Analysis of over 4,000 samples from the biobank revealed a prevalence of anaemia (as per WHO definition haemoglobin less than 11g/dl) of 91% in under-2 year olds and 71% in 2-3 year olds, with 78% of this anaemia (based on MCV cut off 70 fL) derived from well-nourished African American children) caused by iron deficiency (unpublished analysis). Children from the 3 closest villages to the Keneba clinic (Keneba, Kantong Kunda and Manduar) will be recruited when they present for vitamin A supplementation and well-child check-up at 6 months of age, or when self-presenting with an acute infection.

### Recruitment and procedures for IDeA Study 1

#### Study design

This is a prospective cohort study of children who will enroll at the age of 6-8 months, with a follow-up period of one calender year from recruitment onwards. A total of 120 well children will be recruited from weekly vaccination call clinics. After informed consent is obtained, children will have to meet the following inclusion/exclusion criteria to be enrolled.

#### Inclusion/exclusion criteria

Children must be aged between 6-8 months at the time of study enrolment, residing in and intending to stay in the study site area and willing to adhere to all protocol visits and procedures, free from acute illness, with a normal temperature (<37.5 degrees Celsius), with no vaccinations in the last 7 days prior to study enrolment, with no recent administration of immune-suppressants or other immune-modifying agents within 90 days prior to study, with no administration of systemic antibiotic treatment within 3 days prior to study enrolment, with no history of, or evidence for, chronic clinically significant disorder or disease, with no history of maternal infections by human immunodeficiency virus, chronic hepatitis B or chronic hepatitis C, and not participating in another study.

A field worker will be responsible for maintaining links with participants between follow up visits and arranging transport to and from the study site. Scheduling will be arranged through the REDCap software system (REDCap; https://projectredcap.org).

#### Participant flow

We expect to enrol 10 participants per month over one calendar year so that the target sample size (120 children) will be attained in one year. Each participant will be seen three times, at times when they would usually present to clinic as part of The Gambia EPI schedule: 6 months, 12 months (Diptheria, Petussis and Tetanus boosters) and 18 months (Polio, Measles and Rubella boosters). All patients will be seen and take part in the iron absorption test before receiving routine immunisation.

#### Clinical assessment

At each visit, children will be examined by the research clinician using pre-determined checklists for possible sources of low-grade inflammation (**Tables 1**).

**Table 1:** Clinical Score of Site and Severity of Infection and Inflammation (CSSSII)

| System | Sign/Diagnosis | Site | Severity score<br>Normal (0), mild (1), moderate (2), severe (3) |
| --- | --- | --- | --- |
| **Lymphadenopathy** |  | Neck, axilla, groin, other | 0 None (0); 1 1 node < 1cm, rubbery (1); 1-3 nodes, rubbery (2); > 3 nodes, any node > 1cm, any node hard or evidence of suprainfection (3). |
| **Ears** | Otitis externa | Right, left | Normal (0); TM seen, canal erythematous (1); Oedematous canal, debris in canal, TM obscured (2) Fever, oedematous ear canal, cellulitis of pinna (3) |
|  | Otitis media | Right, left | Normal (0); Retracted/neutral position of TM, mild erythema (1), Moderate TM erythema, TM mobility decreased (2), Full, bulging TM, otorrhoea, severe TM erythema, TM mobility absent (3) |
| **Nose** | Discharge |  | None (0); Clear (1); Purulent (2); Profuse, purulent and/ or bloody (3) |
| **Throat** |  | Right, left | Normal (0); Mild inflammation (1); Inflammation and exudate (2); Severe inflammation and swelling exudate/pus/abscess (3) |
| **Respiratory** | Presence of interval symptoms (asthma) |  | No interval symptoms (0); Night time cough, shortness of breath on exertion, or family history, or previous use of inhaler (1); Diagnosed asthma and regular use of inhaler (2) Use of inhalers every day/ active Use of inhalers every day/ active flare up currently |
|  | Oxygen saturations |  | 0 >95% (0); 94-95% (1); 90-93% (2); < 90% (3) |
|  | Tachypnea |  | None (0); Mild intercostal retractions (1); Moderate intercostal retractions (2); Severe, head bobbing, tracheal tugging (3) |
|  | Auscultation |  | No added sounds (0) End expiratory wheeze/ creps only (1) Few scattered/ focal crepitations. Focal reduced air entry (2); Coarse crepitations throughout both lung fields, reduced air entry throughout both lung fields (3) |
| **Skin** | Scabies |  | None (0); < 10 lesions (1); 11-49 lesions (2); > 50 lesions or crusted scabies (3) |
|  | Impetigo |  | None (0)<br>1 1-10 lesions (1)<br>2 11-49 lesions (2)<br>3 > 50 lesions or associated with any systemic symptoms (fever, vomiting, rigors) (3) |
|  | Atopic dermatitis |  | None (0); Less 5% BSA involved, no acute changes, no significant impact on quality of life (1); 5-30% BSA involved, mild dermatitis with acute changes, mild dermatitis with significant impact on Quality of Life (QoL) (2) >30% BSA, Moderate dermatitis with acute changes, Moderate dermatitis with significant impact on Quality of Life (3) |
|  | Oral thrush |  | None (0); White coated tongue, mouth (1); Red painful to eat/swallow (2); Evidence of oesophageal involvement (3) |
|  | Tinea |  | None (0); One plaque (1); 2-5 plaques (2); > 5 plaques (3) |
|  | Nappy Rash |  | None (0); Mild erythema/ papules (1); Broken skin, severe erythema (2); Suprabacterial infection (3) |
|  | Helminths |  | None (0); Larva currens (1); Larva migrans (2); Larva migrans with systemic features (3) |
|  | Trauma |  | None (0); 1-2 bruise or superficial scratch < 5cm diameter (1); single bruise or deeper lacerations 5-10cm or multiple superficial wounds in 2-10 locations (2); Bruise/ laceration >10cm or greater than 10 discrete areas (3) |
| **Eyes** | Infection |  | None (0); Discharge/ mild conjunctivitis in one eye (1); Discharge/ conjunctivitis in both eyes (2); Periorbital or orbital cellulitis (3) |
| **Dental** | Gingivitis |  | None (0); Mild erythema, swelling and bleeding of gums (1); Moderate erythema, swelling, bleeding and signs of infection (2); Severe erythema, swelling, bleeding and infection in gums (3) |
|  | Number of decayed teeth |  | None (0); 1-2 decayed teeth (1); 2-5 (2); 3 >5 (3) |
|  | Mouth and mucous membranes |  | None (0); Angular stomatitis (1); Herpes lesions 1-2 (2) 3 or more herpes lesions or similar (3) |

A clinical score has been devised based on a number of clinical severity scores for certain minor illnesses of childhood and takes into account severity of symptom/condition and the number of anatomical sites affected. Sources of inflammation and infection will be searched for in the ears, nose and throat, the respiratory system, the skin and all skin flexures, the oral cavity including dentition, evidence of lymphadenopathy, signs of infection/inflammation in the orbital region and any evidence of injury or trauma. Severity is graded as 0-normal/no inflammation; 1-mild inflammation; 2-moderate inflammation; 3-severe inflammation. Existing validated clinical scores developed to guide management and treatment were used and adapted for diagnoses of otitis externa [29], otitis media [30], lower respiratory tract infections [31], and atopic dermatitis [32].

Where clinical scores do not exist, divisions of severity will be made based on clinical findings such as number of lesions or extent of erythema. The number of anatomical locations and severity of the signs found is added together to give a numerical score with a range 0-81 (severe inflammation at 27 possible anatomical sites).

Skin conditions, poor oral health, and respiratory infections/airway disease are very common in children in West Kiang, particularly in the rainy season, but are not usually considered serious enough by the parents to bring the child to clinic. Possible skin sources of inflammation include eczema, impetigo, folliculitis, and fungal infections such as tinea capitis will be noted if present. Although gingivitis and dental caries are rare in UK children of this age, poor oral hygiene in the Gambian population Is a potential source of inflammation [33,34].

### Recruitment and procedures for IDeA Study 2

#### Study design

In total, 200 sick children aged between 6-36 months will be recruited from the clinic. Successive children brought by their mothers to the clinic will be recruited into 4 diagnostic categories; upper respiratory tract infections, lower respiratory tract infections, gastroenteritis and urinary tract infections. These reflect the commonest causes of fever in this age range [35,36]. Fever is the commonest presenting complaint to the Keneba clinic in this age range. Each participant will be seen 4 times: Days 0, 3, 7 and 14 after initial presentation. After informed consent is obtained, children will have to meet the following inclusion/exclusion criteria to be enrolled into the study.

#### Inclusion/exclusion criteria

For inclusion, children must be aged between 6-36 months at the time of study enrolment, residing in and intending to stay in the study site area and willing to adhere to all protocol visits and procedures, with a fever (>37.5C) and/or signs of acute illness, with no vaccinations in the last 7 days prior to study enrolment, with no recent administration of immune-suppressants or other immune-modifying agents within 90 days prior to study, with no administration of systemic antibiotic treatment within 3 days prior to study enrolment, with no history of or evidence for chronic clinically significant disorder or disease, with no maternal history of human immunodeficiency virus, chronic hepatitis B or chronic hepatitis C infections, and not participating in another study.

In addition to this they must fall into one of the four following diagnostic categories as defined below.

##### Urinary tract infection (UTI) [37]

Inclusion criteria (one from the list below):

1. Positive leucocytes, positive nitrites on dipstick
2. Negative leucocytes, positive nitrites on dipstick
3. Positive leucocytes, negative nitrites, plus bacteriuria on microscopy
4. Positive leukocytes, negative nitrites plus no bacteriuria, only pyuria on microscopy PLUS clinical features e.g. fever, pain on urination, offensive smelling urine.

##### Lower respiratory tract infections (LRTI) [38]

Inclusion criteria (one from the list below):

1. Focal signs on auscultation of the chest i.e. crepitations/ wheeze/ reduced air entry
2. Fever > 38.5C AND chest recessions AND/OR respiratory rate over 40 breaths/minute
3. Radiological evidence of LRTI

Exclusion criteria:

1. Positive malaria test OR suspicion of metabolic acidosis causing tachypnoea and fever

##### Upper respiratory tract infection (URTI) [39]

Inclusion criteria (one from the list below):

1. Evidence of nasal discharge AND/OR
2. Inflammation throat/ tonsils on direct examination AND/OR
3. Inflammation of middle or outer ear on direct examination
4. History of fever AND history of stridor/ barking cough
5. History of fever AND lymphadenopathy AND/ OR URTI symptoms i.e. sore throat/cough

Exclusion criteria (one from the list below):

1. Foreign body inserted in either nose/ ear
2. Traumatic perforation of ear drum
3. Allergic rhinitis i.e. good contact history
4. Evidence of LRTI

##### Diarrhoea/gastroenteritis [39]

Inclusion criteria (one from the list below):

1. Abrupt onset of 3 or more loose/liquid stools/ day
2. Ova, cysts, parasites identified on stool microscopy PLUS (diarrhoea or fever or vomiting)
3. Fever AND vomiting WITHOUT other cause of fever i.e. UTI/LRTI/URTI

Exclusion criteria (one from the list below):

**1.** Neurological cause of vomiting

###### Withdrawal of participants

In case the participant decides to withdraw during either study, the participant’s samples will not be worked on, but any information previously generated from the samples before the time of withdrawal will be used.

### Oral iron absorption test

After the clinical examination, an oral iron absorption protocol will be initiated in all children:

<u>Step 1:</u> A baseline venous blood sample will be collected for later assay of full blood count (including reticulocyte count) and serum iron markers, hepcidin, erythropoietin (EPO), erythroferrone, IL-6 and EndoCab;
<u>Step 2:</u> Children will be given an oral dose of liquid ferrous fumarate at 2mg/kg;
<u>Step 3:</u> 4 hours later a venous blood sample will be drawn for measurement serum iron markers. The change in serum iron levels (measured before and after dosing) will be used as a direct measurement of iron absorption.

### Laboratory analysis

Whole blood samples will be assessed for: full haematology panel (using a Medonic M20M GP or Cell Dyne Ruby). All serum samples collected will be assessed for the following: hepcidin, erythropoietin, erythroferrone all by ELISA. Additionally, serum ferritin, serum iron, unsaturated iron-binding capacity (UIBC), soluble transferrin receptor (sTfR), transferrin, C-reactive protein (CRP), and alpha 1-acid glycoprotein (AGP) will be assessed using a fully automated biochemistry analyser (Cobas Integra 400 plus).

A stool sample will be collected to check for presence and severity of helminth infection and analysed for fecal calprotectin as a marker of gut inflammation or environmental enteric dysfunction (EED).

### Data entry, handling, storage and security

All protocol-required field data will be captured electronically on an electronic Case Report Form (eCRF). Data will be entered in real time using eCRFs developed in a REDCap (Research Electronic Data Capture) database. Entered data will be acquired by the database via a direct secure connection over the 4G mobile network. Laboratory related data will be extracted directly from laboratory equipment, where functionality is available and uploaded to the database. Otherwise, results will be captured using a custom designed laboratory results form. Any data collected on the paper format will be double entered by a trained data entry clerk.

After giving written consent, the children will be given a study identification number, which will be used in all future datasets for subject anonymity. Any and all paper/electronic forms will be checked and any errors will be identified prior to marking data as complete. Electronic data will be stored on the local dedicated server maintained at MRCG. The study will be conducted in compliance with Good Clinical Practice. Study personal security measures will include controlled access limited to authorised users only, physical security, removal of identifiable information (anonymization) prior to any data sharing, avoidance of third-party cloud storage and password protection.

### Sample size calculation

The primary objective is a descriptive study. No sample size calculation was conducted.

### Statistical analysis

#### Primary objective

The primary objective of this study is to determine the sources of low-grade inflammation in otherwise-well children between the ages of 6-24 months living in rural Gambia.

#### Analysis plan

Multiple regression analysis will be used to model inflammatory markers (serum concentrations of CRP and AGP) as a function of clinical scores for skin infections, oral health and respiratory infections; and stool markers of inflammation (fecal calprotectin). In these analyses, we will account for child age as a potential confounder, and we will account for multiple assessments for each child.

#### Secondary objective 1

To retest the hypothesis that falling hepcidin concentrations predict a rise in serum iron absorption in a much larger sample size.

#### Analysis plan

Multiple regression analysis will be used to model iron absorption (serum iron concentration at 3h after oral administration of the iron supplement) as a function of hepcidin concentration. Because we expect the serum iron response to strongly depend on serum iron concentration before administration of the iron dose), we will include initial serum iron concentration as a covariate in the model (both as main term and its product term with hepcidin concentration). We will also assess the role of other potential effect modifiers (age, anaemia, CRP, AGP, sTfR, MCV, EPO and erythroferrone), each in separate models, using a backwards elimination process.

#### Secondary objective 2

To establish the role of inflammation in the relationship between EPO and haematocrit to test for decreased EPO synthesis and/or increased EPO resistance in anaemic children with low-grade inflammation.

#### Analysis plan

In the absence of anaemia, EPO concentrations are low (approximately 10U/L) and independent of haemoglobin concentration, consistent with our biological understanding that EPO is produced only in response to hypoxic stress. In anaemia, however, EPO production may increase by a factor of up to 1000, with the increase in log EPO being linearly associated with haemoglobin concentration (or its proxy, haematocrit values). The slope of the regression line between log EPO and haemoglobin concentration is reduced in patients with inflammation compared to their peers with moderate to severe deficit of functional iron. In iron deficiency without inflammation, it is believed that there are no restrictions in EPO production other than its response to hypoxia. Together, this evidence indicates that, in inflammation, the EPO response is inappropriately low for the degree of anaemia.

The challenge in our analysis will be to similarly show the influence of inflammation (indicated by serum concentrations of CRP or AGP) on the association between EPO and haemoglobin concentration. Because the degree of inflammation is expected to be lower than in the patients studied in previously published reports, and because haemoglobin concentration is expected to be higher, we expect much smaller effects.

We will assess influences on EPO using fractional polynomial regression models with CRP and AGP (continuous variables) and haemoglobin concentration (continuous variable centred at 110g/L), as well as their product terms, whilst accounting for dependency of observations within persons and potential heteroscedasticity:

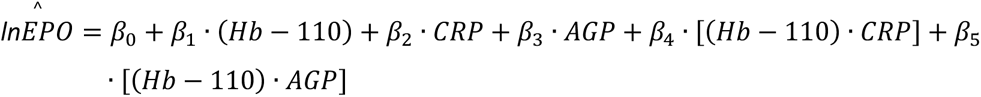

where Hb indicates haemoglobin concentration in g/L. Analysis will be restricted to individuals with anaemia (haemoglobin concentration ≤110g/L). We expect to be able to eliminate the main terms for CRP and AGP from this model, under the assumptions that inflammation does not lead to a reduction in EPO concentration unless it is accompanied by a reduction in haemoglobin concentration below 110g/L, and that inflammation is adequately and entirely represented by CRP and AGP. In such conditions, inflammation will influence the slope of the line between log EPO and haemoglobin concentration, but not its intercept (i.e., at haemoglobin concentrations of 110g/L, our model above will reduce to a single expected value: 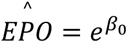). Thus, our model is parametrised to allow evaluation of our hypothesis that inflammation reduces EPO response to haemoglobin concentration (*β*_4_ and *β*_5_ are both below zero).

In an analysis of all data combined, the potential effect modification by CRP (and alternate measures of iron status and inflammation including hepcidin) will be assessed using forward selection to determine the variables for inclusion in multivariate analysis of factors predicting the slope and intercept of the EPO/Hb relationship. We will also calculate the Reticulocyte Production Index (RPI) = (Hct/45)*Retic/Maturation. An RPI >3 shows a normal marrow response to anemia. An RPI <2 is an inadequate response to anemia and is consistent with a diagnosis of AI. Additionally, because serum sTfR levels are the product of cellular receptor density (and are up-regulated by iron demand) and cell numbers (primarily deemed to reflect erythroblasts) we will conduct an exploratory analysis of the inter-relationships between EPO, sTfR and reticulocyte number.

#### Secondary objectives 3 and 4

To describe the interrelationships between: a) systemic inflammation (CRP/AGP) and its associations with hepcidin and ERFE, and b) the influence of EPO on the association between Hb and ERFE.

#### Analysis plan

We will conduct a hypothesis-free exploratory analysis to assess whether ERFE behaves as predicted based by mouse models (ie up-regulated by stress erythropoiesis and inversely related to hepcidin). Our first task will be to map and explore the inter-relationships between ERFE and the erythroid drive (reticulocytes, sTfR and EPO) and to examine whether there is evidence that ERFE is significantly modulating hepcidin. This will require multiple regression modelling and pathways analysis that include the competing stimuli for which prior analysis suggest that UIBC and MCV will be the best indices of iron status and CRP/AGP and granulocyte counts the best indices of infection/inflammation. Depending upon the outcomes of these exploratory analyses we may be able to test our hypothesis that the high levels of hepcidin so common in Gambian children are driven in part by suppressed ERFE.

### Confounding, bias and loss to follow up

The study design does not provide for the recruitment of equal numbers of subjects in each month of the year (or during the dry vs wet seasons). If we are missing data for the covariates, for the secondary analysis, we will use pairwise deletion and consider multiple imputation if there are missing covariates in more than 5% of participants and data can be assumed to be missing at random. If loss to follow-up rate is considerably different between groups, we will perform sensitivity analyses to examine the robustness of results.

## DISCUSSION

Iron deficiency and iron deficiency anaemia remain significant public health concerns in children in The Gambia and similar LMICs. Programmes to combat iron deficiency are frequently unsuccessful due to poor efficacy of oral iron supplementation and potential hazards in certain circumstances. We have previously shown that the low-grade inflammation frequently found in children living in unsanitary environments can result in hepcidin-mediated blockade of iron absorption. This project will try to identify the sources of this low-grade inflammation in seemingly ‘well’ Gambian children and assess its severity in unwell children. It will additionally try to understand the complex interacting mechanisms linking inflammation, hepcidin, EPO and ERFE to iron absorption, distribution and erythropoiesis. If, as we speculate, the origins of IDA are more strongly determined by inflammation than by low dietary iron this would have important implications for future clinical interventions as well as agricultural and livestock policies.

## Data Availability

All data produced in the present study are available upon reasonable request to the authors

## Abbreviations

AGP: alpha-1-acid-glycoprotein
CRP: C-reactive protein
ELISA: enzyme-linked immunosorbent assay
EndoCAB: serum endotoxin core antibody
EPO: erythropoietin
ERFE: erythroferrone
Hb: haemoglobin
ID: iron deficiency
IDA: iron deficiency anaemia
iFABP: enterocyte fatty acid binding protein
IL-6: interleukin 6
IRIDA: iron refractory iron deficiency anaemia
LMIC: lower middle income country
MCV: mean cell volume
REG1b: regenerating protein 1 beta
RPI: retriculocyte production index
ROC AUC: area under the receiver operating characteristic curve
sTfR: soluble transferrin receptor
UIBC: unsaturated iron-binding capacity
WHO: World Health Organisation.

## Acknowledgments

We thank the Dr Fatai Akemokwe and Edrisa Sinjanka for their guidance and enthusiasm during the planning this study.

## Availability of data and materials

All data will be made available to researchers upon reasonable request to the study PI and clearance by the MRCG Scientific Coordinating and Ethics Committees.

## Authors contributions

AMP and CC conceived the study. EL and CC designed the clinical study protocol. EL, CC, ATJ developed the laboratory protocols. HV wrote the data analysis plan in consultation with CC and AMP. EL, NC, CC and AMP drafted the manuscript. All authors approved the manuscript.

## Ethics approval and consent to participate

These studies have been approved by The Gambia Government/MRC Joint Ethics Committee (LEO 1664 and 17079). The study procedures will be explained to the children’s mothers/guardians orally with a witness present if they are illiterate or in writing. A child will be recruited into the study after the consent form has been signed/thumb printed by the mother/guardian.

### Future use of stored specimens

The blood and faecal samples collected during the trial may be used to support other research in the future, and may be shared anonymously with other researchers, for their ethically approved projects. Any future use would require approvals from the principal investigator, MRCG@LSHTM Scientific Coordinating Committee, and the Gambian Government MRCG Joint Ethics Committee.

Informed consent from the parents/guardians for this will be included within the informed consent documents.

### Participant confidentiality

Any identifiable data collected will be stored securely and their confidentiality protected in accordance with the UK General Data Protection Regulation (UK GDPR) and the Data Protection Act (2018). All data will be anonymised, and individuals will not be identifiable.

### Dissemination and data availability

All key findings from this study will be submitted for publication in peer-reviewed journals. Any request for use of study data will go through approval from the Scientific Coordinating Committee at MRC Unit The Gambian and the Gambian Government MRCG Joint Ethics Committee. All data will be in an anonymous format for external users.

## Consent for publication

Not applicable.

## References

1. James SL et al. Global, regional, and national incidence, prevalence, and years lived with disability for 354 diseases and injuries for 195 countries and territories, 1990–2017: a systematic analysis for the Global Burden of Disease Study 2017. The Lancet. 2018;392:1789–1858.

2. Hess SY, McLain AC, Frongillo EA, Afshin A, Kassebaum NJ, Osendarp SJM, Atkin R, Rawat R, Brown KH. Challenges for Estimating the Global Prevalence of Micronutrient Deficiencies and Related Disease Burden: A Case Study of the Global Burden of Disease Study. Curr Dev Nutr. 2021;5:nzab141.

3. Cusick SE & Georgieff MK. The Role of Nutrition in Brain Development: The Golden Opportunity of the “First 1000 Days.” J Pediat. 2016;175:16–21.

4. Georgieff MK, Ramel SE, Cusick SE. Nutritional influences on brain development. Acta Paediatr. 2018;107:1310–1321.

5. Horton S, & Ross J. The economics of iron deficiency. Food Policy. 2003;28:51–75.

6. Drakesmith H, et al. Vaccine efficacy and iron deficiency: an intertwined pair? Lancet Haematol. 2021;8:e666–e669.

7. Drakesmith H & Prentice AM. Hepcidin and the iron-infection axis. Science. 2012;338:768–72.

8. Prentice AM, Mendoza YA, Pereira D, Cerami C, Wegmuller R, Constable A & Spieldenner J. Dietary strategies for improving iron status: Balancing safety and efficacy. Nutrition 2017;75:49–60.

9. Clark MA et al. Host iron status and iron supplementation mediate susceptibility to erythrocytic stage plasmodium falciparum. Nat Comms. 2014;5:4446

10. Goheen MM, et al. Anemia Offers Stronger Protection Than Sickle Cell Trait Against the Erythrocytic Stage of Falciparum Malaria and This Protection Is Reversed by Iron Supplementation. EBioMedicine. 2016;14:123–130

11. Cross JH, Bradbury RS, Fulford AJ, Jallow AT, Wegmüller R, Prentice AM & Cerami C. Oral iron acutely elevates bacterial growth in human serum. Scientific Reports. 2015;5:16670

12. Bah A, et al. Hepcidin-guided screen-and-treat interventions against iron-deficiency anaemia in pregnancy: a randomised controlled trial in The Gambia. Lancet Glob Health. 2019;7:e1564–e1574.

13. Wegmüller R et al. Efficacy and safety of hepcidin-based screen-and-treat approaches using two different doses versus a standard universal approach of iron supplementation in young children in rural Gambia: A double-blind randomised controlled trial. BMC Pediatrics. 2016;16:149.

14. Wegmüller R, Bah A, Kendall L, Goheen MM, Sanyang S, Danso E, Sise EA, Jallow A, Verhoef H, Jallow MW, Wathuo M, Armitage AE, Drakesmith H, Pasricha SR, Cross JH, Cerami C, Prentice AM. Hepcidin-guided screen-and-treat interventions for young children with iron-deficiency anaemia in The Gambia: an individually randomised, three-arm, double-blind, controlled, proof-of-concept, non-inferiority trial. Lancet Glob Health. 2023;11:e105–e116.

15. Tam E, Keats EC, Rind F, Das JK, Bhutta AZA. Micronutrient Supplementation and Fortification Interventions on Health and Development Outcomes among Children Under-Five in Low- and Middle-Income Countries: A Systematic Review and Meta-Analysis. Nutrients. 2020;12:289.

16. Ganz T. Systemic iron homeostasis. Physiol Rev. 2013;93:1721–41.

17. Weiss G, Ganz T, Goodnough LT. Anemia of inflammation. Blood. 2019;3:133:40–50.

18. Prentice AM et al. Respiratory infections drive hepcidin-mediated blockade of iron absorption leading to iron deficiency anemia in African children. Science Advances. 2019;5:eaav9020

19. Doherty CP, Cox SE, Fulford AJ, Austin S, Hilmers DC, Abrams SA, Prentice AM. Iron incorporation and post-malaria anaemia. PLoS One. 2008;3:e2133.

20. Prentice AM, et al. Hepcidin is the major predictor of erythrocyte iron incorporation in anemic African children. Blood. 2012;119:1922–8.

21. de Mast Q, et al. Assessment of urinary concentrations of hepcidin provides novel insight into disturbances in iron homeostasis during malarial infection. J Infect Dis. 2009 199:253–62.

22. Cusick SE, Opoka RO, Abrams SA, John CC, Georgieff MK, Mupere E. Delaying Iron Therapy until 28 Days after Antimalarial Treatment Is Associated with Greater Iron Incorporation and Equivalent Hematologic Recovery after 56 Days in Children: A Randomized Controlled Trial. J Nutr. 2016;146:1769–74.

23. Pasricha SR et al. Expression of the iron hormone hepcidin distinguishes different types of anemia in African children. Sci Trans Med. 2014;6:235re3.

24. van der Vorm LN et al. Toward worldwide hepcidin assay harmonization: Identification of a commutable secondary reference material. Clin Chem. 2016;62:993–1001.

25. Kurtzhals JA, Rodrigues O, Addae M, Commey JO, Nkrumah FK, et al. Reversible suppression of bone marrow response to erythropoietin in Plasmodium falciparum malaria. Br J Haematol 1997;97:169–174.

26. Kautz L, Jung G, Valore EV, Rivella S, Nemeth E & Ganz T. Identification of erythroferrone as an erythroid regulator of iron metabolism. Nat Genet. 2014;46:678–84

27. Pasricha S-R, McHugh K & Drakesmith H. Regulation of Hepcidin by Erythropoiesis: The Story So Far. Ann Rev Nutr. 2016;36:417–34

28. Hennig BJ, et al. Cohort Profile: The Kiang West Longitudinal Population Study (KWLPS)-a platform for integrated research and health care provision in rural Gambia. Int J Epidemiol. 2012;46:e13

29. Hajioff D & Mackeith S. Otitis externa. BMJ Clin Evid. 2015;2015:0510

30. Casey JR, Block S, Puthoor P, Hedrick J, Almudevar A & Pichichero ME. A simple scoring system to improve clinical assessment of acute otitis media. Clin Ped. 2011;50:623–9.

31. Rodriguez H, Hartert TV, Gebretsadik T, Carroll KN & Larkin EK. A simple respiratory severity score that may be used in evaluation of acute respiratory infection Pediatrics. BMC Research Notes. 2016;9:85.

32. Wolkerstorfer A, De Waard van der Spek FB, Glazenburg EJ, Mulder PGH & Oranje AP. Scoring the severity of atopic dermatitis: Three item severity score as a rough system for daily practice and as a pre-screening tool for studies. Acta Dermato-Venereologica. 1999;79:356–9.

33. Gordon N. Oral health care for children attending a malnutrition clinic in South Africa. Int J Dent Hyg. 2007;5:180–6

34. Lamb WH & Prentice AM. A prospective survey of gingivitis in Keneba, a rural West African community. Annals of Tropical Paediatrics. 1983;3:137–42.

35. D’Acremont V et al. Beyond malaria - Causes of fever in outpatient Tanzanian children. New Engl J Med 2014;370:809–17.

36. Davis T. NICE guideline: Feverish illness in children - Assessment and initial management in children younger than 5 years. Arch Dis Childh: Educ Pract Ed. 2013;98:232–5.

37. NICE. (NICE) Urinary tract infection in under 16s Diagnosis and management. National Institute for Health and Care Excellence (2007).

38. Harris M et al. British Thoracic Society guidelines for the management of community acquired pneumonia in children: Update 2011. Thorax 2011;66 Suppl 2:ii1–23

39. World Health Organization. Guideline: Updates on Paediatric Emergency Triage, Assessment and Treatment: Care of Critically-Ill Children. Guideline: Updates on Paediatric Emergency Triage, Assessment and Treatment: Care of Critically-Ill Children (2016).

